# Reporting results of diagnostic NGS-based testing – user-centered redesign of the clinical report

**DOI:** 10.1101/2021.03.15.20197236

**Authors:** Oleg Agafonov, Sigrun Vik, Kaja O. Kjølås, Sharmini Alagaratnam

## Abstract

The transition of next-generation sequencing (NGS) from the research environment to clinical diagnostics has proven difficult, with exome and whole genome sequencing at various stages of implementation in Nordic hospitals. The clinical genomics report details key findings from the interpretation of NGS data and represents the core hand-off between specialized clinical genomics laboratories and the broader healthcare community. However, these text-heavy reports can be difficult to read: critical information may be scattered around the report, and vital information such as limitations of the test may not always be present. Misunderstanding of results, limitations or key findings can lead to incorrect therapeutic decisions, directly impacting patient management.

We applied the principles of user-centred design to redesign clinical reports to respond to user needs, while also incorporating existing recommendations and guidelines. We performed several rounds of needs gathering, first with producers of clinical genetics reports, then with clinicians with various level of experience in genetic testing. Based on the insights from a workshop and interviews, we created prototypes of reports which were evaluated in comparison to a set of simulated reports representing those currently in clinical production. Our results showed that the majority of evaluators found the redesigned reports to be clearer and easier to process, demonstrating the value of this approach.

## 2 Introduction

Next-generation sequencing (NGS) has been widely used for research purposes for over 15 years since the first generation of massive parallel sequencers appeared on the market (1), followed by its application in routine diagnostics. Nevertheless, the transition from the research environment to clinical diagnostics has proven difficult. NGS diagnostics for rare diseases is a complex procedure comprising multiple steps – collection and DNA extraction from a biological sample such as blood, then DNA sequencing and bioinformatic analysis, resulting in a lengthy list of genetic variants. On average a human genome differs from the reference genome at 4.1 million to 5.0 million sites (2), therefore a series of filtering steps are required to narrow down the search to variants potentially associated with a disease phenotype, before final manual interpretation of the candidate variants. Once a conclusion is reached, the diagnostic laboratory describes the result of the test in the clinical genomics report, along with information on how this result was obtained. The goal of this report is both to transfer this sophisticated information from diagnostic laboratories to the clinician managing the patient, and provide guidance for subsequent clinical decision making (3).

The complexity of NGS-based diagnostic test result originates in the inherent uncertainties of the testing procedure - only variants located in genes or non-coding parts of the genome deemed relevant for a specific disease phenotype are usually subjected to analysis; and only certain types of genetic variants are currently analysed, namely single nucleotide variants (SNVs), small insertions and deletions and copy number variants (CNVs), while other types (e.g. complex structural variants) are often excluded from the analysis today. Overall, the diagnostic yield of NGS-based diagnostic tests varies from 7 to 42%, depending on the type of test and condition, the availability of family members for analysis and the type of genetic variant (4–7). When identified, the reported molecular variant is assigned probability as a cause of the disease phenotype, reflecting the level of certainty (8,9). In contrast to many other types of tests, the absence of findings of a molecular cause of the disease phenotype does not necessarily imply the absence of disease, resulting in limited negative predictive value. In addition to pathogenic findings, some laboratories report variants of uncertain significance (VUS), a class of variants that do not have sufficient evidence to be classified as benign or pathogenic (10), but may have significance for the management of disease with future scientific discoveries (11–13), and should therefore be followed up on. Taken together, all these factors underline the challenges of transferring often subtle but critical diagnostic information. As a result, the topic of clinical reporting was nominated as one of the key challenges in NGS-based diagnostics by members of clinical genetics laboratories at a biannual Nordic Alliance for Clinical Genomics workshop (14).

In our previous study, we reviewed existing guidelines for clinical reporting and identified recommendations available in the scientific literature spanning various parts of the reporting process and report content (15). Among these recommendations, four areas were identified as especially challenging to implement: VUS, secondary findings, reanalysis and data delivery to patients. We also conducted a peer-review benchmarking of Nordic reports form NSG-based diagnostic tests where we provided fictional clinical cases to diagnostic laboratories and asked them to produce reports for these, which then were evaluated by specialists in medical genetics and NGS-based diagnostic testing. Through this benchmarking exercise, we found that even though evaluators scored the reports as high in clarity, they experienced difficulties in identifying the presence of key information in the report, for example if secondary findings were systematically searched for. This exercise indicated that even though laboratories were aware of the challenges surrounding clinical reporting, there is room for improvement in practice; in particular difficulties in comprehension of key information were identified as being due not only to report design and formatting, which can be limited by local IT production systems, but also methods for delivering the report to the requisitioning clinician (15).

Clinical reports currently in use in the Nordics have evolved along with the development of testing technologies and pre-existing IT report production infrastructure, and do not necessarily respond to user needs. In this work, we applied the principles of user-centred design (UCD) to redesign clinical reports according to user needs, while also incorporating existing recommendations and legislation identified in our previous study (15). UCD is an approach that incorporates the user’s perspective into the product development process to achieve an adequate and usable product (16). This approach has been internationally accepted and defined as an ISO standard - ISO 9241-210 (17). The standard states six principles which should be considered for product development:

- product development should be based on users and their tasks and esnvironments
- end-users should be involved in product design and development
- product development should be based on input from end-users
- the product development process should be iterative
- the design should address the whole user experience
- the design team should be multidisciplinary.

In the redesign process, we identified a range of potential report users through a workshop with clinical genetics laboratories from five Nordic countries: Denmark, Finland, Iceland, Norway and Sweden. To identify and analyze user needs for clinical reports in medical genetics, we conducted semi-structured interviews with clinicians and report producers. Taking into consideration the needs identified, we created prototype clinical reports utilizing a multidisciplinary team of designers and specialists in medical genetics and NGS diagnostics. Finally, we tested the prototypes with end-users by benchmarking the prototypes against simulated existing reports, identifying opportunities for iterative improvement.

## 3 Methods

### 3.1 UNDERSTANDING THE CONTEXT OF USE AND IDENTIFYING USER NEEDS

#### 3.1.1 Exploratory kick-off workshop with report producers

This workshop took place during the Nordic Alliance for Clinical Genomics meeting in Copenhagen in November 2018. Eighteen clinicians and diagnostic laboratory members from 5 Nordic countries participated in the workshop, during which main groups of clinical report users, existing activities and challenges were identified.

#### 3.1.2 Semi-structured interviews with clinicians and patients

##### Sampling

Convenience and snowball sampling were used leveraging the internal and external networks of the research team and the Nordic Alliance for Clinical Genomics to recruit clinicians and NGS diagnostic laboratories staff for interviews.

##### Data collection

Semi-structured interviews were focused on exploring interviewees’ perspectives based on their experiences of receiving reports. Interviews lasted between 45 to 90 minutes and were conducted between February and April 2019. List of interview questions can be found in Appendix 1, Table 1.

**Table 1.**
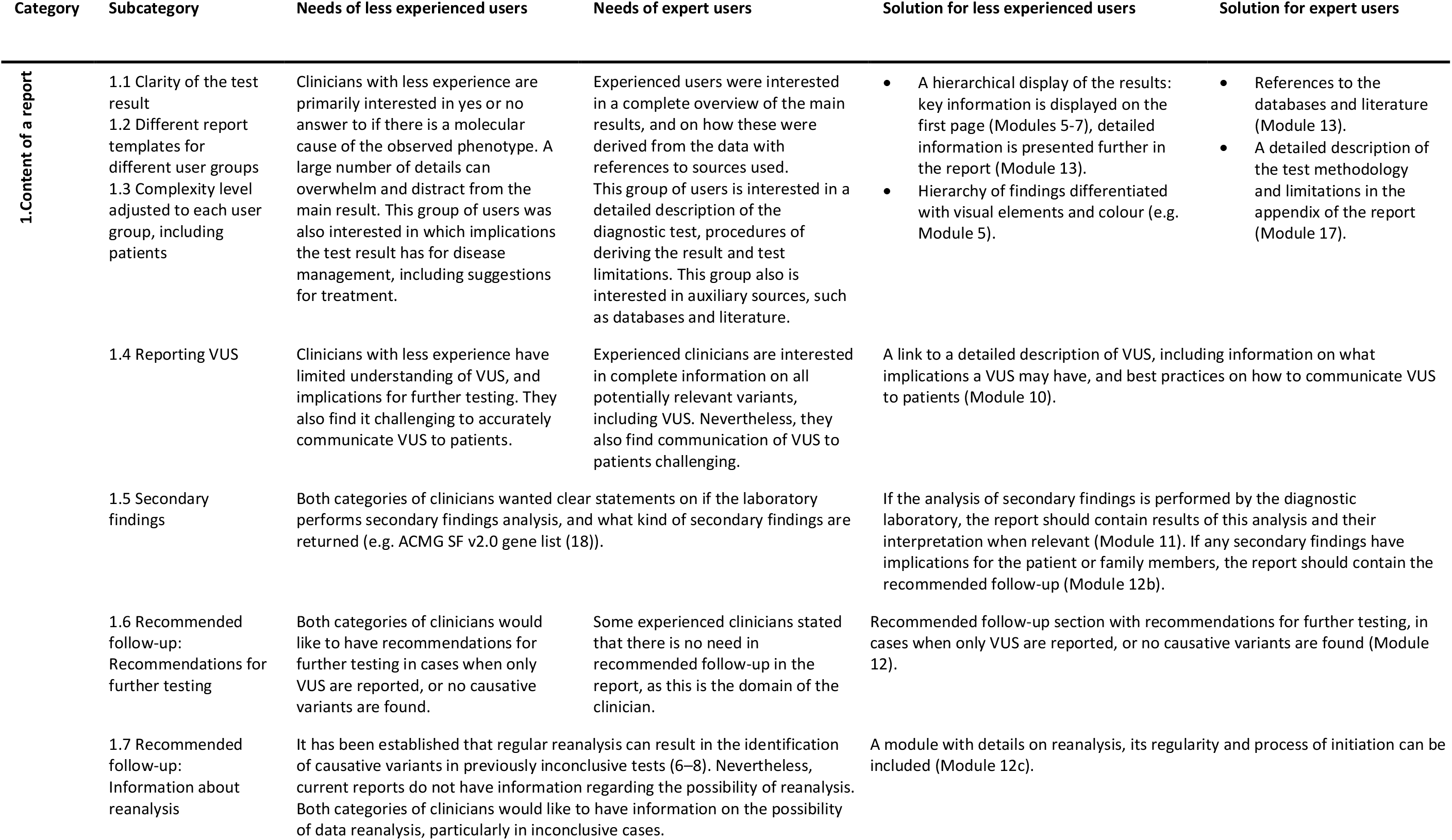
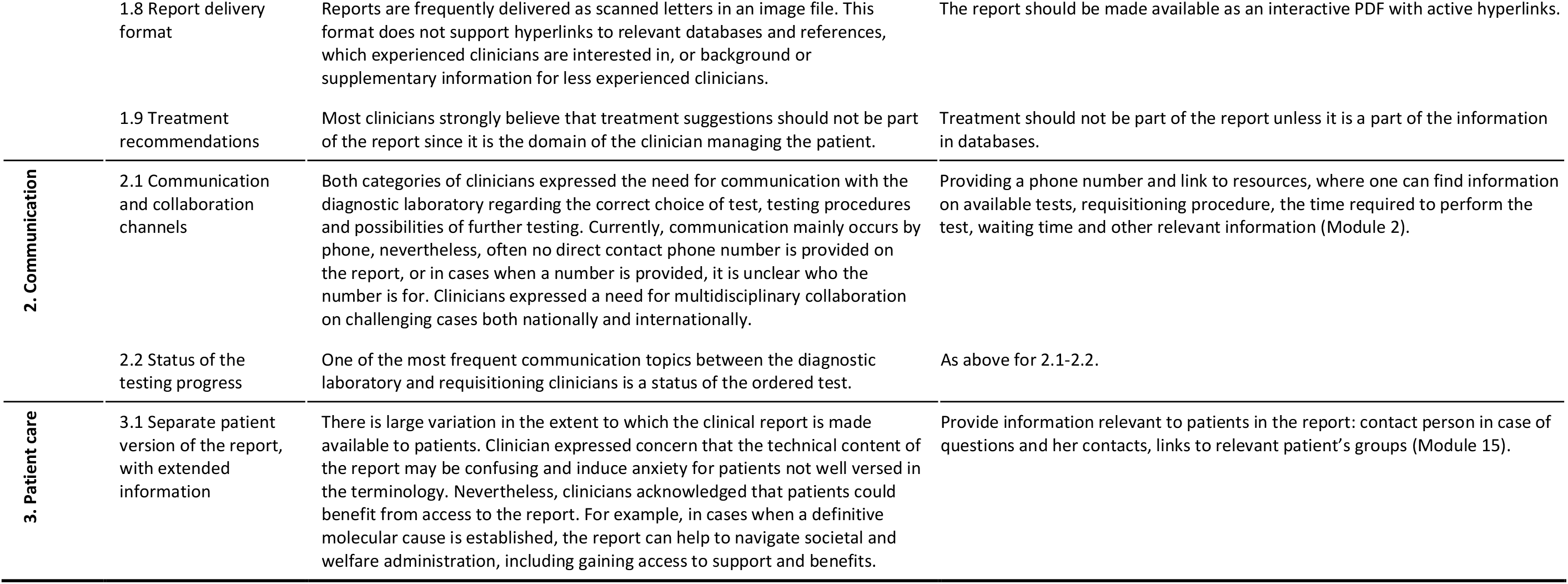
User needs and proposed design solutions

##### Data analysis

The interview responses (n=9) were collated for content analysis. Significant words or phrases were extracted from interview notes. The words and phrases with similar meanings were categorized together and organized into common themes. Taking an iterative approach, a series of workshops were held among the researchers to discuss and condense the themes and their descriptions. Team consensus was used to resolve any disagreement.

##### Ethical considerations

Interviewees participated voluntarily and all interviewees gave their consent verbally. Interview responses were anonymized and treated as highly confidential. Only the research team had access to the responses. In terms of GDPR compliance, a clear project policy was developed of not receiving or recording any personal information, and communicated to all project members.

### 3.2 PRODUCING PROTOTYPE REPORTS

Interviews were analysed individually by the project team consisting of both design and clinical genetics experts to extract challenges related to the process of information transfer in clinical NGS reporting. Storyboarding was used to capture the key moments of the user journey into stories to support visual ideas. The insights gathered through this were grouped according to themes, and ideation methods were used to visualize potential solutions responses to these needs. Wireframes, or quick, low-fidelity sketches, were first developed to address the main concepts of layout and usability. Design modules for combinatorial use were refined. Finally, prototype and prototype elements were identified and prioritized on the modular components, in collaboration with clinicians and researchers.

### 3.3 EVALUATING THE PROTOTYPE REPORTS

A survey for report producers and users was designed to validate the redesigned clinical reports prototypes and test the accuracy of data flow from producer to user, and user satisfaction. Each participant received two sets of reports: the first set simulated current reports and contained the main elements of a clinical NGS report, highlighted with colour and structured with tables. We used a simulation, rather than in-production reports for benchmarking to avoid familiarity bias. The second set contained the redesigned prototype reports (see Appendix 2). Both sets contained three fictional clinical cases: the first with a likely pathogenic finding, the second with a VUS and a secondary pathogenic finding in the *BRCA2* gene, and the third with no findings of clinical significance. Participants, clinicians with various level of familiarity with NGS-based testing (n=8) were asked to answer a series of questions for each report, where the survey contained Likert scale questions, ‘Yes’ or ‘no’ questions and open-ended questions (see Appendix 1, Appendix Table 2). Surveys allowed for additional qualitative feedback on topics discussed.

## 4 Results

### 4.1 REDESIGN PROCESS OVERVIEW

To redesign clinical reports, we utilized UCD methodology by first conducting a workshop with report producers to identify the full spectrum of report users, main challenges in clinical reporting, and to conceptualize ideal solutions. We then conducted a series of semi-structured interviews (n=9) with report users: clinicians with various levels of familiarity with genetic testing, various frequency of communication with diagnostic laboratories and of various ages. After that, we developed discrete report modules, where each module addressed a specific aspect of a report (e.g. patient information, results, conclusion, etc.) which could be modified separately and combined in various layouts. These modules were used to produce prototypes that were tested on a small number of clinicians and report producers; and insights from testing used to modify the report’s structure and content. Finally, the resulting prototypes were tested in comparison to existing reporting solutions on a broader scope of clinicians; see Figure 1.

**Figure 1.**
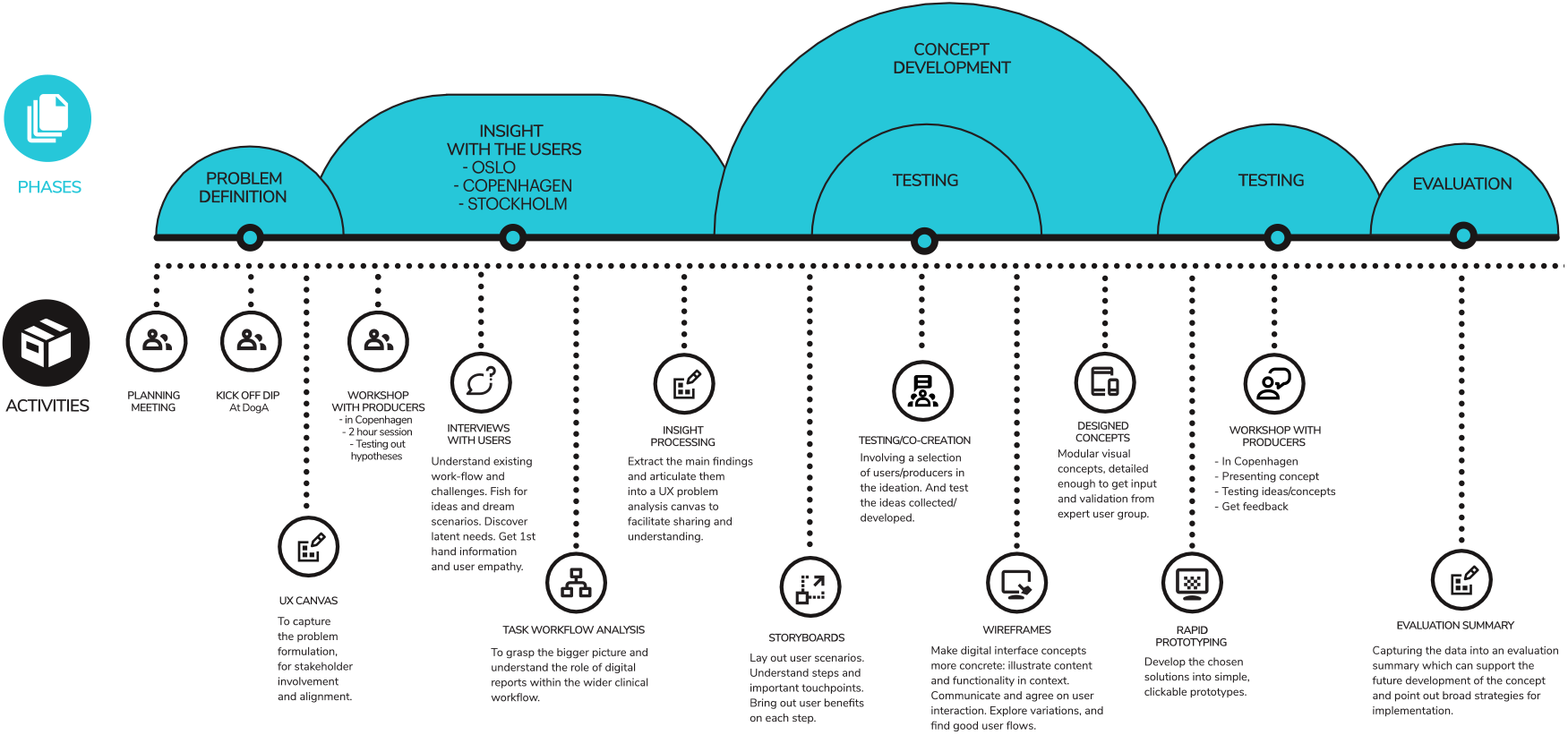
Process map of the redesign.

### 4.2 UNDERSTANDING AND SPECIFYING THE CONTEXT OF THE USE

#### 4.2.1 The users and their characteristics

In an earlier study, we found wide variation among clinical professionals in how NGS-based testing reports are read and processed (15). To sufficiently address user needs, we needed to identify their characteristics. In a workshop with clinical genetics laboratories from five Nordic countries, we were able to identify several categories of clinical report users. The first consists of requisitioning clinicians, which is a diverse group of health personnel, ranging from general practitioners who only occasionally requisition genetic testing (several cases per year, we refer to this group as clinicians with less experience in genetic testing) to clinical specialists who requisition genetic testing regularly (several cases per week, we refer to this group as clinicians with a high level of competence in genetics), e.g. psychiatrists and neurologists at regional centres. The second group consists of producers of the report, laboratory personnel at hospitals who perform NGS testing. The third group of users are patients and patients’ legal guardians. Although there was variation in how often clinicians provided clinical reports to their patients, some patients specifically requested these reports. In some cases, patients or patients’ legal guardians may provide clinical reports to patient organizations for support and guidance. Clinicians also reported anecdotal cases of patients uploading their clinical reports to social networks/forums to discuss diagnosis, making them available for the general public.

In the workshop, producers were asked to nominate specific clinicians with a range of genetic literacy, ages and frequency of requisitioning clinical NGS tests for in-depth semi-structured interviews. Report producers also proposed their perceived needs and potential challenges of reports’ users, which we used to create a questionnaire for the semi-structured interviews to identify user needs.

#### 4.2.2 Processing content of a report by different groups of users

To investigate how the reports are used by clinicians, we conducted a series of semi-structured interviews. Questions addressed the process of initiating testing, receiving a report, processing its content, communication with the diagnostic laboratory and patients, and potential ideal solutions.

##### Variation in the way reports are perceived

We found significant variation in the way reports are read and processed by different categories of clinicians. Clinicians with a high level of competence in genetics were interested in receiving a detailed report to enable as complete as possible understanding of the diagnostics laboratory’s findings. This group preferred reports without recommendations for the treatment (which is the current practice), believing this to be within their domain of responsibility. Clinicians with less experience in genetic testing were primarily interested in a clear test result (for example in an easy-to-understand “traffic light” representation), with specific suggestions for follow-up and/or disease management.

##### Variants of uncertain significance

Uncertain test results were particularly challenging for clinicians with less experience in genetic testing. VUS are variants that do not have sufficient evidence to be classified as pathogenic or benign, and some laboratories report these variants to ensure that requisitioning clinicians receive a complete overview of all variants. Clinicians with less experience in genetic testing had difficulties in understanding the relevance of such variants to the disease phenotype. Clinicians regularly dealing with NGS-based testing stated that they had no problem understanding the uncertain result, but also stated that communicating uncertain results to patients was challenging.

##### Negative test result

Similarly to uncertain findings, understanding the implications of a negative result was also challenging for clinicians with less experience in genetic testing, since it required a detailed understanding of the performed test, including its limitations.

##### Delivery of reports to patients

Although anecdotal evidence showed that some clinicians believed patients and/or their next-of-kin would not understand details of an NGS-based diagnostic report, they acknowledged that receiving a report with a clear diagnosis could help patients or patients next-of-kin to navigate societal structures and organizations to secure support for themselves or their child.

#### 4.2.3 The environment of the report

Clinical reports were found to be most frequently delivered either as a paper letter, which is then scanned and attached to the patient’s electronic health record, or directly as a scanned document. In both cases, the final document can often only be accessed as an image file, which does not allow the use of active hyperlinks to relevant sources of information such as databases or additional details regarding the testing procedure, or copying text. Several clinicians expressed concern that they needed to retype Human Genome Variation Society (HGVS) nomenclature terms or other technical details to search for additional information regarding reported variants, in a time-consuming and potentially error-prone process. In some cases, the report is scanned by administrative personnel, and the requisitioning clinician may not be notified about the received report, introducing additional delays to the patient receiving a diagnosis.

### 4.3 SPECIFYING THE USER NEEDS AND PRODUCING DESIGN SOLUTIONS

We collected user needs by interviewing report producers and users, and during the workshop with report producers. As a result, we identified areas that should be addressed in the redesign process of the report, summarized in

Table 1. To create a report prototype we utilized a modular design approach, where each module addresses specific tasks and user needs, Figure 2.

**Figure 2.**
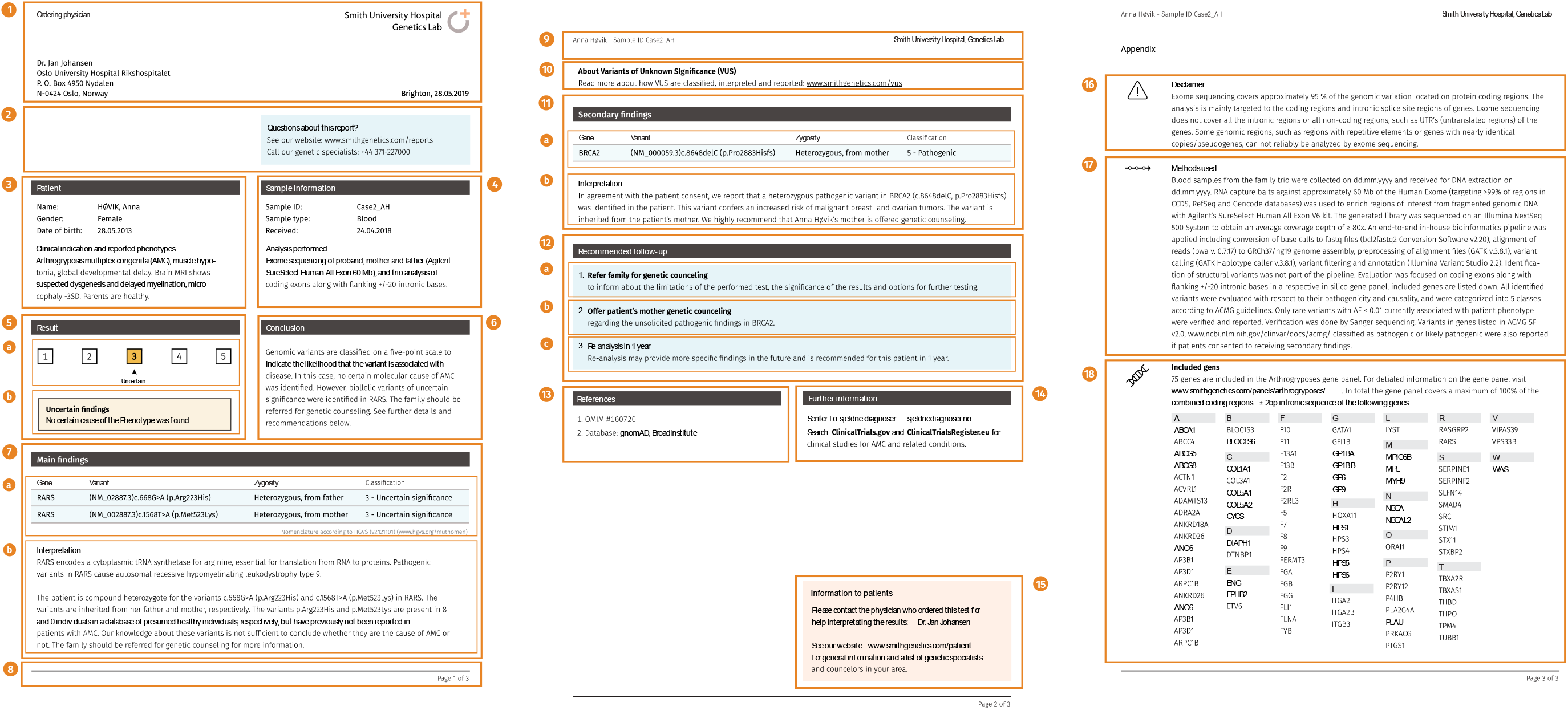
Example of a redesigned clinical report. 1 – header with information on requisitioning clinician and diagnostic laboratory; 2 – contact information with phone number and a web-link to laboratory information site; 3 – patient details, including description of phenotype; 4 – information on sample; 5 – main result, including colour-coded category; 6 – summary of test conclusion; 7 – main findings (a) and their interpretation (b); 8 – footer with number of page number and total number of pages; 9 – header with patient name and sample ID; 10 – supplementary information on VUS (present only in reports that contain VUS); 11 – secondary findings (a) details and (b) interpretation (present only in reports that contain secondary findings); 12 – recommended follow-up with recommendations on genetic counselling (a,b), recommended reanalysis (c); 13 – relevant references; 14 – further information with links to relevant clinical trials and other relevant information; 15 – information for patients including details on who to contact in case of questions, and relevant links with information on genetic testing, patients groups, etc.; 16 – disclaimer stating test limitations; 17 – methodology of genetic testing; 18 – list of tested genes (or in cases when the list is too long, a link to this list).

### 4.4 EVALUATING THE DESIGN

During the evaluation, we aimed to test if the redesigned reports performed better in comparison to the simulation of an existing report. Simulated reports were created based on the combination of the production reports of participating institutions – Oslo University Hospital in Norway, Rigshospitalet in Denmark, and HUSLAB in Finland. During the report simulation, we attempted to create a report which resembles reports from our partners instead of using existing actual partner reports to avoid familiarity bias during the evaluation.

During the evaluation, participants were asked a series of questions addressing various parts of the reports; see Figure 3. In addition, we recorded and analysed comments made by evaluators.

**Figure 3.**
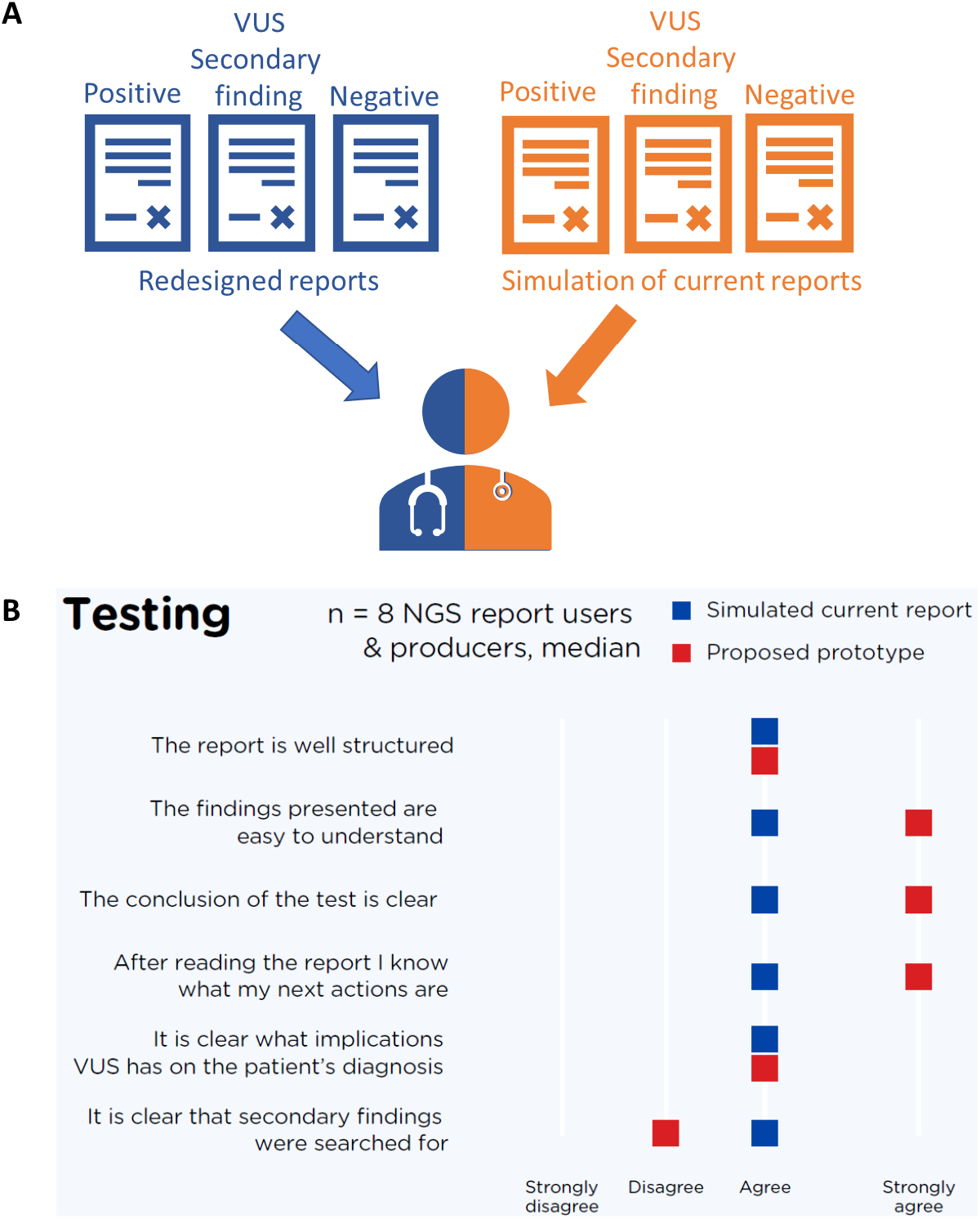
Validation of redesigned reports with a survey. A. Validation test design. B Summary of the survey result for Likert scale questions.

#### Visual appearance

Participants found redesigned reports more visually attractive and easier to navigate, stating the structure to be clearer and more intuitive. In particular, evaluators found that a clear representation of the main result, including a clear indication of the class of reported variants (Module 5), made it easier to read the report.

#### Recommended follow-up

Evaluators noted that recommended follow-up section (Module 12) is clear and useful, however, some expressed concern that opinion of a clinician responsible for the patient may differ from the opinion of the laboratory stated in the recommended follow-up, which may cause conflict in patient management.

#### Reanalysis policy

Evaluators expressed a preference for information regarding reanalysis policy (Module 12c) to be present in all reports, while in redesigned reports it was available only in reports without findings and reports with VUS.

#### References

Evaluators noted that references to relevant scientific publications and databases (Module 13) were helpful when additional information is needed for assessing the patient.

#### Supporting information

Evaluators stressed the importance of the appendix with supporting information, including methodology, limitations (Modules 16-17) and list of genes (Module 18), stating that this should always be present in the report.

#### Secondary findings

Some evaluators stated that it was not clear to them if secondary findings were present in the redesigned reports, and this information should be more clearly presented. A potential solution could be a module on a first page of the report, clearly stating presence or absence of secondary findings.

## Discussion

In this work, we utilised a user-centred design methodology to redesign an NGS-based clinical test report. Through the process of insight gathering from report users, we better understood the cause of variation on how a clinical report is processed by various groups of users. Through this, we were also better placed to address the main challenge of redesigning this report: that the same report is delivered to and must respond to different groups of users with different levels of genetic literacy and needs. The main strategy applied to overcome this challenge was to apply a clear hierarchy in the modular visualization of the results in the report, highlighting the key results of the test and recommended follow-up for the clinicians who are most interested in these. At the same time, more detailed information such as references to relevant literature sources and databases were made available for clinicians who were keen to access and process this information, as well as a detailed description of the methodology, test limitations, and list of tested genes being specified in the report’s appendix.

A clinical report with a confirmed diagnosis can act as important documentation for patients to access support and welfare benefits, and both according to our survey and as reported earlier, patients do in fact often receive clinical reports (19). Despite this, and the impact of a confirmed diagnosis or the lack thereof being greatest for the patient, current versions of the reports of the laboratories that participated in our study lack supporting information for understanding the report for patients and/or their next of kin. Ideally, patients should have access to genetic counselling to answer their questions related to their diagnosis. However, patients may have additional questions at various stages, particularly in the case of uncertain or no findings. Some laboratories provide a separate report for patients (19), nevertheless this approach burdens laboratories with additional work, and is not scalable with the increasing number of genetic tests performed unless a higher degree of automation is used in report production. In the redesigned report prototype we attempt to address patients’ needs by including a contact number, links to relevant sources of information about genetic testing, their disease and relevant patient support groups. This gives the patient recourse to contact information, answers to their initial questions, and navigates to trustworthy sources of information.

The clinical report is an important and tangible deliverable during the process of diagnostic NGS testing, yet continuous communication between the diagnostic laboratory and requisitioning clinicians is critical for the final outcome. Clinicians who requisitioned genetic testing more frequently reported a lower threshold for contacting the diagnostic laboratory with questions regarding an appropriate choice of test than clinicians who requisitioned genetic testing more rarely. At the same time, diagnostic laboratories reported that clinicians who requisitioned genetic testing more rarely are less likely to provide all the information necessary for the most appropriate test to be chosen. Similarly, upon receiving the clinical report, expert users are more comfortable and more frequently contact the diagnostic laboratory for additional information when needed than less experienced requisitioners. We positioned contact information for the diagnostic laboratory strategically in the redesigned reports and used language intended to lower the threshold for any reader of the report to take contact in case of any questions, and generally to encourage communication.

In this work, we attempted to create prototype reports responding to user needs that can be implemented within a reasonable timeframe. Despite this, the barriers represented by rigid IT infrastructure and production systems for implementing design solutions should not be underestimated, where anecdotal evidence shows instances where only plain text can be entered to report the result of a diagnostic test. However, with the evolution of IT infrastructure over time, there is room for further improvement of the delivery of clinical reports, and indeed overall communication and interaction between requisitioning clinicians, diagnostic laboratories and patients, from assistance with test requisition, status of the testing process and dialogue after the test result is ready.

## Conclusion

The clinical NGS report serves to transmit the results of this diagnostic test to requisitioning clinicians who have variable knowledge and level of experience with such tests, leading to wide variation in how the report is read and perceived. Complex information needs to be presented clearly, yet without oversimplification to the point of potentially misguiding clinical decisions. Many current reporting solutions have not been designed per se, but rather represent inherited properties of reports for other types of tests, and genetic testing laboratories often lack resources and in-house UX competence to design a report. The reports delivered by commercial providers of genetic tests or analysis tend to be an exception, nevertheless these reports also vary in comprehensiveness and usability. Here we have attempted to meet user needs by creating clinical NGS report prototypes with differentiated levels of information comprising of individual modules, which can be entirely or flexibly integrated with existing reporting systems used in clinics. The created report prototype is distributed with a [Creative Commons knowledge license](https://creativecommons.org/licenses/by-nc/4.0/), and can be freely used for non-commercial purposes.

## Data Availability

No supplementary data is available

## Acknowledgements

The research was conducted with the support of Design and Architecture Norway, grant number 295393. The authors thank clinicians and members of clinical genetic laboratories for participating in interviews, workshops and surveys. The authors thank Morten C. Eike, Bobbie Nicole Ray-Sannerud, Tita Alissa Bach and Courtney David Nadeau for reviewing the manuscript draft, Kaja Kvello and Tita Alissa Bach for assisting with organization, execution and processing of interviews and workshops.

## Appendixes

Appendix 1. Interviews questions.

Appendix 2. Redesigned reports for the three fictitious cases used for testing.

