## Appendix for "Reporting results of diagnostic NGS-based testing – user-centered redesign of the clinical report"

### APPENDIX 1

**Appendix Table 1.** List of interview questions to collect information on how a report is processed by different groups of users.

| Theme | Questions |
|---|---|
| Content/layout | How do you receive your reports and what format are they in?<br>In what order are you looking for information in the report?<br>What do you want to know first?<br>Where to find it?<br>Are there specific concepts or points you are looking for when scanning through the report?<br>What is the essential information?<br>What is hard to understand/easy to understand?<br>How do you know which analyses are done and not done, and why is it important?<br>How often do reports inform treatment directly?<br>How do you know what the next step in the process is?<br>Do you know what is expected of you?<br>What do you like/dislike about reports? |
| Communication | With the patient:<br>• How and what do you communicate to your patients to communicate the results?<br>• Do you refer patients for genetic counselling?<br>With the producer:<br>• How often do you have to go back to NGS lab for clarification? |
| The dream | What challenges and needs are there from your perspective?<br>If the clinical report was not a paper report/PDF, what would it be instead? |

**Appendix Table 2**. List of interview questions for evaluation of developed prototypes.

| Theme | Questions |
|---|---|
| Content/layout | It is easy to find result of the test in the report (Scale)<br>The information in the report is complete (Scale)<br>The results/findings presented in the report are easy to understand (Scale)<br>The report is well-structured (Scale)<br>The conclusion of the test is clear (Scale)<br>After reading the report I know what my next step is (Scale) |
| VUS | It is clear what implications VUS has on the patient's diagnosis (Scale) |
| Secondary findings | It is clear that secondary findings were intentionally searched for (Scale)<br>Please indicate if secondary findings were included in the report (Yes/No) |
| Data reanalysis | Does the report contain information about the possibility of future data reanalysis? (Yes/No) |
| Communication | It is easy to use the report to communicate findings to a patient (Scale) |
| General open-ended questions | Which set of reports do you prefer? Why? (open-ended)<br>What did you like best in the report template? (open-ended)<br>What did you like least in the report template? (open-ended)<br>Do you have any suggestions to improve the template? (open-ended) |

### APPENDIX 2

A set of redesigned prototype reports for three fictional clinical cases: report A contains a likely pathogenic finding; report B contains a VUS and a secondary pathogenic finding in the BRCA2 gene; report C contains no findings of clinical significance.

Smith University Hospital
Genetics Lab

Dr. Jan Johansen
Smith University Hospital
Garry Building, Western Rd,
Brighton BN3 5BW, UK

Brighton, 28.05.2019

**Questions about this report?**
See our website: www.smithgenetics.com/reports
Call our genetic specialists: +44 371-227000

#### Patient

| | |
|---|---|
| Name: | NORDAHL, Mattias |
| Gender: | Male |
| Date of birth: | 22.08.2018 |

**Clinical indication and reported phenotypes**
Diagnosis arthrogryposis multiplex congenita (AMC). Contractures in fingers, feet and knees. Big skull with prominent forehead with hemangioma that spreads over thoracic spine and sacral pit. Short nose, long filtrum, downward mungipor, long eyelashes.

#### Sample information

| | |
|---|---|
| Sample ID: | Case1_MN |
| Sample type: | Blood |
| Received: | 20.05.2019 |

**Analysis performed**
Exome sequencing of proband, mother and father (Agilent SureSelect Human All Exon 60 Mb), and trio analysis of coding exons along with flanking +/-20 intronic bases.

#### Result

1 2 3 **4** 5

**Likely pathogenic**

**Pathogenic findings**
A likely pathogenic variant in MYH3 was found.

#### Conclusion

Genomic variants are classified on a five-point scale to indicate the likelihood that the variant is associated with disease. In this case, a likely pathogenic variant in MYH3 was identified. We interpret this as the likely cause of the patient's phenotype and as genetic evidence of the diagnosis Arthrogryposis Multiplex Congenita (AMC)[1].

We highly recommend that the family is referred for genetic counseling as follow-up. See further details and recommendations below.

#### Main findings

| Gene | Variant | Zygosity | Classification |
|---|---|---|---|
| MYH3 | (NM_002470.3)c.700G>A (p.Ala234Thr) | Homozygous | 4 - Likely pathogenic |

Nomenclature according to HGVS (v2.121101) (www.hgvs.org/mutnomen)

**Interpretation**
A biallelic variant in MYH3 was identified in the patient. MYH3 encodes the embryonic isoform of myosin heavy-chain (MyHC) that is expressed during fetal life. Myosin is a molecular motor and the essential part of the thick filament of striated muscle.

Pathogenic variants in MYH3 are reported to cause autosomal dominant distal arthrogryposis type 2A, 2B and 8 and autosomal recessive spondylocarpotarsal synostosis syndrome.

**Continued on next page**

The patient is homozygous for c.700G>A (p.Ala234Thr) in MYH3, inherited from heterozygous parents. The variant is not present in a database of presumed healthy individuals[2] and it is reported heterozygote in two individuals with distal arthrogryposis. We regard this variant as being likely pathogenic and thus interpret this to be the likely cause of the patient's phenotype, and as genetic evidence of the diagnosis Arthrogryposis Multiplex Congenita (AMC).

We recommend that the family is referred for genetic counseling for follow-up.

#### Recommended follow-up

1. **Refer family for genetic counseling**
   Inform of the consequences of the findings in this report and discuss options for further testing.
2. **Annual screening for liver disease**
   Pathogenic variants in MYH can be associated with an increase risk of liver disease, annual screening is recommended.

#### References

1. OMIM #618484
2. Database: gnomAD, Broadinstitute

#### Further information

Foreningen for Muskelsyke: **ffm.no**

Senter for sjeldne diagnoser: **sjeldnediagnoser.no**

Search **ClinicalTrials.gov** and **ClinicalTrialsRegister.eu** for clinical studies on AMC and related conditions.

##### Information for patients

Please contact the physician who ordered this test for help interpreting the results: **Dr. Jan Johansen**

See our website **www.smithgenetics.com/patient** for general information and a list of genetic specialists and counselors in your area.

**Continued on next page**

### Appendix

#### Disclaimer

Exome sequencing covers approximately 95 % of the genomic variation located on protein coding regions. The analysis is mainly targeted to the coding regions and intronic splice site regions of genes in the insilico gene panel (see list of included genes below). Exome sequencing does not cover all intronic regions or all non-coding regions, such as UTR's (untranslated regions) of genes. Some genomic regions, such as regions with repetitive elements or genes with nearly identical copies/pseudogenes, can not reliably be analyzed by exome sequencing.

#### Methods used

Blood samples from the family trio were collected in EDTA tubes 20.05.2019 and received for sequencing on the same day. Genomic DNA was extracted with Agilent gDNA Extraction Kit following the manufacturer's protocol. The exome library was captured with the Agilent SureSelect Human All Exon V6 kit following the manufacturer's protocol. The generated library was sequenced on an Illumina NextSeq 500 System to an average coverage depth of ≥ 80x.

An end-to-end in-house bioinformatics pipeline was applied including conversion of base calls to fastq files (bcl2fastq2 Conversion Software v2.20), alignment of reads (bwa v. 0.7.17) to GRCh37/hg19 genome assembly, pre-processing of alignment files (GATK v.3.8.1), variant calling (GATK Haplotype caller v.3.8.1), variant filtering and annotation with Illumina Variant Studio 2.2. The bioinformatics pipeline called SNPs and INDELs shorter than 50 bp.

Evaluation was limited to the HCM LQT insilico gene panel (see details below), and ACMG recommended genes for reporting of secondary findings, www.ncbi.nlm.nih.gov/clinvar/docs/acmg/ (Kalia et al., 2017) with a focus on coding exons along with flanking +/-20 intronic bases. Variants were categorized into 5 classes according to the ACMG guidelines (Richards et al., 2015). Only rare variants with allele frequency lower than 0.01 currently associated with patient phenotype were verified by Sanger sequencing and reported. Variants classified as pathogenic, likely pathogenic and variants of uncertain significance are included in the report.

#### Included genes

75 genes are included in the insilico HCM LQT gene panel. For detailed information on the gene panel visit **www.smithgenetics.com/panels/HCM_LQT/**. In total the gene panel HCM LQT covers a maximum of 100% of the combined coding regions ± 2bp intronic sequence of the following genes:

**A**
ABCA1
ABCC4
ABCG5
ABCG8
ACTN1
ACVRL1
ADAMTS13
ADRA2A
ANKRD18A
ANKRD26
ANO6
AP3B1
AP3D1
ARPC1B
ANKRD26
ANO6
AP3B1
AP3D1
ARPC1B

**B**
BLOC1S3
BLOC1S6

**C**
COL1A1
COL3A1
COL5A1
COL5A2
CYCS

**D**
DIAPH1
DTNBP1

**E**
ENG
EPHB2
ETV6

**F**
F10
F11
F13A1
F13B
F2
F2R
F2RL3
F5
F7
F8
F9
FERMT3
FGA
FGB
FGG
FLI1
FLNA
FYB

**G**
GATA1
GFI1B
GP1BA
GP1BB
GP6
GP9

**H**
HOXA11
HPS1
HPS3
HPS4
HPS5
HPS6

**I**
ITGA2
ITGA2B
ITGB3

**L**
LYST

**M**
MPIG6B
MPL
MYH3

**N**
NBEA
NBEAL2

**O**
ORAI1

**P**
P2RY1
P2RY12
P4HB
PLA2G4A
PLAU
PRKACG
PTGS1

**R**
RASGRP2
RUNX1

**S**
SERPINE1
SERPINF2
SLFN14
SMAD4
SRC
STIM1
STX11
STXBP2

**T**
TBXA2R
TBXAS1
THBD
THPO
TPM4
TUBB1

**V**
VIPAS39
VPS33B

**W**
WAS

B

Ordering physician

Smith University Hospital
Genetics Lab

Dr. Jan Johansen
Smith University Hospital
Garry Building, Western Rd,
Brighton BN3 5BW, UK

Brighton, 28.05.2019

**Questions about this report?**
See our website: www.smithgenetics.com/reports
Call our genetic specialists: +44 371-227000

#### Patient

Name: HOVIK, Anna
Gender: Female
Date of birth: 28.05.2013

##### Clinical indication and reported phenotypes

Arthrogryposis multiplex congenita (AMC), muscle hypotonia, global developmental delay. Brain MRI shows suspected dysgenesis and delayed myelination, microcephaly -3SD. Parents are healthy.

#### Sample information

Sample ID: Case2_AH
Sample type: Blood
Received: 24.04.2018

##### Analysis performed

Exome sequencing of proband, mother and father (Agilent SureSelect Human All Exon 60 Mb), and trio analysis of coding exons along with flanking +/-20 intronic bases.

#### Result

1 2 **3** 4 5

Uncertain

**Uncertain findings**
No certain cause of the phenotype was found.

#### Conclusion

Genomic variants are classified on a five-point scale to indicate the likelihood that the variant is associated with disease. In this case, no certain molecular cause of AMC[1] was identified. However, biallelic variants of uncertain significance were identified in RARS. The family should be referred for genetic counseling. See further details and recommendations below.

#### Main findings

| Gene | Variant | Zygosity | Classification |
|---|---|---|---|
| RARS | (NM_02887.3)c.668G>A (p.Arg223His) | Heterozygous, from father | 3 - Uncertain significance |
| RARS | (NM_002887.3)c.1568T>A (p.Met523Lys) | Heterozygous, from mother | 3 - Uncertain significance |

Nomenclature according to HGVS (v2.121101) (www.hgvs.org/mutnomen)

##### Interpretation

RARS encodes a cytoplasmic tRNA synthetase for arginine, essential for translation from RNA to proteins. Pathogenic variants in RARS cause autosomal recessive hypomyelinating leukodystrophy type 9.

The patient is compound heterozygous for the variants c.668G>A (p.Arg223His) and c.1568T>A (p.Met523Lys) in RARS. The variants are inherited from her father and mother respectively. The variants p.Arg223His and p.Met523Lys are present in 8 and 0 individuals in a database of presumed healthy individuals[2] respectively, but have not previously been reported in patients with AMC. Our knowledge about these variants is not sufficient to conclude whether they are the cause of AMC or not. The family should be referred for genetic counseling for more information.

**Continued on next page**

**About Variants of Unknown SIgnificance (VUS)**
Read more about how VUS are classified, interpreted and reported: www.smithgenetics.com/vus

#### Secondary findings

| Gene | Variant | Zygosity | Classification |
|---|---|---|---|
| BRCA2 | (NM_000059.3)c.8648delC (p.Pro2883Hisfs) | Heterozygous, from mother | 5 - Pathogenic |

**Interpretation**
In agreement with patient consent, we report that a heterozygous pathogenic variant in BRCA2 (c.8648delC, p.Pro2883Hisfs) was identified in the patient. This variant confers an increased risk of malignant breast- and ovarian tumors. The variant is inherited from the patient's mother. We highly recommend that Anna Høvik's mother is offered genetic counseling.

#### Recommended follow-up

1. **Refer family for genetic counceling**
   to inform about the limitations of the performed test, the significance of the results and options for further testing.
2. **Offer patient's mother genetic counceling**
   regarding the secondary findings in BRCA2.
3. **Re-analysis in 1 year**
   Re-analysis may provide more specific findings in the future and is recommended for this patient in 1 year.

#### References

1. OMIM #160720
2. Database: gnomAD, Broadinstitute

#### Further information

Senter for sjeldne diagnoser: **sjeldnediagnoser.no**

Search **ClinicalTrials.gov** and **ClinicalTrialsRegister.eu** for clinical studies for AMC and related conditions.

##### Information for patients

Please contact the physician who ordered this test for help interpretating the results: **Dr. Jan Johansen**

See our website **www.smithgenetics.com/patient** for general information and a list of genetic specialists and councelors in your area.

**Continued on next page**

#### Appendix

##### Disclaimer

Exome sequencing covers approximately 95 % of the genomic variation located on protein coding regions. The analysis is mainly targeted to the coding regions and intronic splice site regions of genes in the insilico gene panel (see list of included genes below). Exome sequencing does not cover all intronic regions or all non-coding regions, such as UTR's (untranslated regions) of genes. Some genomic regions, such as regions with repetitive elements or genes with nearly identical copies/pseudogenes, can not reliably be analyzed by exome sequencing.

##### Methods used

Blood samples from the family trio were collected in EDTA tubes 18.04.2018 and received for sequencing on the same day. Genomic DNA was extracted with the Agilent gDNA Extraction Kit following the manufacturer's protocol. The exome library was captured with the Agilent SureSelect Human All Exon V6 kit following the manufacturer's protocol. The generated library was sequenced on an Illumina NextSeq 500 System to an average coverage depth of ≥ 80x.

An end-to-end in-house bioinformatics pipeline was applied including conversion of base calls to fastq files (bcl2fastq2 Conversion Software v2.20), alignment of reads (bwa v. 0.7.17) to GRCh37/hg19 genome assembly, pre-processing of alignment files (GATK v.3.8.1), variant calling (GATK Haplotype caller v.3.8.1), variant filtering and annotation with Illumina Variant Studio 2.2. The bioinformatics pipeline called SNPs and INDELs shorter than 50 bp.

Evaluation was limited to the Arthrogryposes insilico gene panel (see details below), and ACMG recommended genes for reporting of secondary findings, www.ncbi.nlm.nih.gov/clinvar/docs/acmg/ (Kalia et al., 2017) with a focus on coding exons along with flanking +/-20 intronic bases. Variants were categorized into 5 classes according to the ACMG guidelines (Richards et al., 2015). Only rare variants with allele frequency lower than 0.01 currently associated with patient phenotype were verified by Sanger sequencing and reported. Variants classified as pathogenic, likely pathogenic and variants of uncertain significance are included in the report.

##### Included genes

75 genes are included in the insilico Arthrogryposes gene panel. For detailed information on the gene panel visit **www.smithgenetics.com/panels/arthrogryposes/**. In total the gene panel covers a maximum of 100% of the combined coding regions ± 2bp intronic sequence of the following genes:

**A**
ABCA1
ABCC4
ABCG5
ABCG8
ACTN1
ACVRL1
ADAMTS13
ADRA2A
ANKRD18A
ANKRD26
ANO6
AP3B1
AP3D1
ARPC1B
ANKRD26
ANO6
AP3B1
AP3D1
ARPC1B

**B**
BLOC1S3
BLOC1S6

**C**
COL1A1
COL3A1
COL5A1
COL5A2
CYCS

**D**
DIAPH1
DTNBP1

**E**
ENG
EPHB2
ETV6

**F**
F10
F11
F13A1
F13B
F2
F2R
F2RL3
F5
F7
F8
F9
FERMT3
FGA
FGB
FGG
FLI1
FLNA
FYB

**G**
GATA1
GFI1B
GP1BA
GP1BB
GP6
GP9

**H**
HOXA11
HPS1
HPS3
HPS4
HPS5
HPS6

**I**
ITGA2
ITGA2B
ITGB3

**L**
LYST

**M**
MPIG6B
MPL
MYH9

**N**
NBEA
NBEAL2

**O**
ORAI1

**P**
P2RY1
P2RY12
P4HB
PLA2G4A
PLAU
PRKACG
PTGS1

**R**
RASGRP2
RARS

**S**
SERPINE1
SERPINF2
SLFN14
SMAD4
SRC
STIM1
STX11
STXBP2

**T**
TBXA2R
TBXAS1
THBD
THPO
TPM4
TUBB1

**V**
VIPAS39
VPS33B

**W**
WAS

C

Ordering physician

Smith University Hospital
Genetics Lab

Dr. Jan Johansen
Smith University Hospital
Garry Building, Western Rd,
Brighton BN3 5BW, UK

Brighton, 28.05.2019

**Questions about this report?**
See our website: http://www.smithgenetics.com/reports
Call our genetic specialists: +44 371-227000

#### Patient

| | |
|---|---|
| Name: | CARLSON, Ivar |
| Gender: | Male |
| Date of birth: | 28.01.1989 |

**Clinical indication and reported phenotypes**
Dyskinetic cerebral palsy.
Epilepsy, scoliosis, speech disorder.

#### Sample information

| | |
|---|---|
| Sample ID: | Case3_IC |
| Sample type: | Blood |
| Received: | 21.05.2019 |

**Analysis performed**
Exome sequencing of proband, mother and father (Agilent SureSelect Human All Exon 60 Mb), and trio analysis of coding exons along with flanking +/-20 intronic bases.

#### Result

**No certain cause of the phenotype was found.**

#### Conclusion

No pathogenic variants are reported, and no certain molecular cause of dyskinetic cerebral palsy was identified in the patient.

#### Recommended follow-up

1. **Refer family for genetic counceling**
   to inform about the limitations of the performed test, the significance of the results and options for further testing.
2. **Re-analysis in 1 year**
   Re-analysis may provide more specific findings in the future and is recommended for this patient in 1 year.

##### Information for patients

Please contact the physician who ordered this test for help interpretating the results: **Dr. Jan Johansen**

See our website **www.smithgenetics.com/patient** for general information and a list of genetic specialists and councelors in your area.

**Continued on next page**

### Appendix

#### Disclaimer

Exome sequencing covers approximately 95 % of the genomic variation located on protein coding regions. The analysis is mainly targeted to the coding regions and intronic splice site regions of genes in the insilico gene panel (see list of included genes below). Exome sequencing does not cover all intronic regions or all non-coding regions, such as UTR's (untranslated regions) of genes. Some genomic regions, such as regions with repetitive elements or genes with nearly identical copies/pseudogenes, can not reliably be analyzed by exome sequencing.

#### Methods used

Blood samples from the family trio were collected in EDTA tubes 21.05.2019 and received for sequencing on the same day. Genomic DNA was extracted with the Agilent gDNA Extraction Kit following the manufacturer's protocol. The exome library was captured with Agilent SureSelect Human All Exon V6 kit following the manufacturer's protocol. The generated library was sequenced on an Illumina NextSeq 500 System to an average coverage depth of ≥ 80x.

An end-to-end in-house bioinformatics pipeline was applied including conversion of base calls to fastq files (bcl2fastq2 Conversion Software v2.20), alignment of reads (bwa v. 0.7.17) to GRCh37/hg19 genome assembly, pre-processing of alignment files (GATK v.3.8.1), variant calling (GATK Haplotype caller v.3.8.1), variant filtering and annotation with Illumina Variant Studio 2.2. The bioinformatics pipeline called SNPs and INDELs shorter than 50 bp.

Evaluation was limited to the Epilepsy insilico gene panel (see details below), and ACMG recommended genes for reporting of secondary findings, www.ncbi.nlm.nih.gov/clinvar/docs/acmg/ (Kalia et al., 2017) with a focus on coding exons along with flanking +/-20 intronic bases. Variants were categorized into 5 classes according to the ACMG guidelines (Richards et al., 2015). Only rare variants with allele frequency lower than 0.01 currently associated with patient phenotype were verified by Sanger sequencing and reported. Variants classified as pathogenic, likely pathogenic and variants of uncertain significance are included in the report.

#### Included genes

281 genes are included in the insilico Epilepsy gene panel. For detailed information on the gene panel visit **www.smithgenetics.com/panels/epilepsy/**. In total the gene panel covers a maximum of 100% of the combined coding regions ± 2bp intronic sequence of the following genes:

**A**
ABCA1
ABCC4
ABCG5
ABCG8
ACTN1
ACVRL1
ADAMTS13
ADRA2A
ANKRD18A
ANKRD26
ANO6
AP3B1
AP3D1
ARPC1B
ANKRD26
ANO6
AP3B1
AP3D1
ARPC1B

**B**
BLOC1S3
BLOC1S6

**C**
COL1A1
COL3A1
COL5A1
COL5A2
CYCS

**D**
DIAPH1
DTNBP1

**E**
ENG
EPHB2
ETV6

**F**
F10
F11
F13A1
F13B
F2
F2R
F2RL3
F5
F7
F8
F9
FERMT3
FGA
FGB
FGG
FLI1
FLNA
FYB

**G**
GATA1
GFI1B
GP1BA
GP1BB
GP6
GP9

**H**
HOXA11
HPS1
HPS3
HPS4
HPS5
HPS6

**I**
ITGA2
ITGA2B
ITGB3

**L**
LYST

**M**
MPIG6B
MPL
MYH9

**N**
NBEA
NBEAL2

**O**
ORAI1

**P**
P2RY1
P2RY12
P4HB
PLA2G4A
PLAU
PRKACG
PTGS1

**R**
RASGRP2
RUNX1

**S**
SERPINE1
SERPINF2
SLFN14
SMAD4
SRC
STIM1
STX11
STXBP2

**T**
TBXA2R
TBXAS1
THBD
THPO
TPM4
TUBB1

**V**
VIPAS39
VPS33B

**W**
WAS
WDR26
WDR45
WWOX

**Y**
YY1

**Z**
ZEB2
ZFYVE26
